# “My friends would believe my word”: appropriateness and acceptability of respondent-driven sampling in recruiting young tertiary student men who have sex with men for HIV/STI research in Nairobi, Kenya

**DOI:** 10.1101/2021.10.21.21265250

**Authors:** Samuel Waweru Mwaniki, Peter Mwenda Kaberia, Peter Mwangi Mugo, Thesla Palanee-Phillips

**Affiliations:** School of Clinical Medicine, Faculty of Health Sciences, University of the Witwatersrand, Johannesburg, South Africa; University Health Services, University of Nairobi, Nairobi, Kenya; School of Mathematics, College of Biological and Physical Sciences, University of Nairobi, Nairobi, Kenya; Kenya Medical Research Institute - Wellcome Trust Research Programme, Nairobi, Kenya; Wits Reproductive Health and HIV Institute, Faculty of Health Sciences, University of the Witwatersrand, Johannesburg, South Africa

**Author notes:** Corresponding author (SWM).

## Abstract

**Background:** Tertiary student men who have sex with men (TSMSM) may engage in behaviors that increase their risk of infection with human immunodeficiency virus (HIV) and sexually transmitted infections (STI). Respondent-driven sampling (RDS) has become a popular method for discretely recruiting marginalized populations into HIV/STI research. We conducted formative research to assess appropriateness and acceptability of RDS in recruiting TSMSM into a prospective HIV/STI bio-behavioral survey in Nairobi, Kenya.

**Methods:** Between September and October 2020, semi-structured qualitative interviews were held with service providers from organizations that serve MSM (n=3), and TSMSM (n=13). Interviews explored social networks of TSMSM, acceptability of using RDS as a sampling method, potential RDS implementation challenges, and proposed solutions to these challenges. Interviews were done in English, audio-recorded and transcribed then analyzed thematically using NVivo version 11.

**Results:** Service providers reflected that TSMSM had large though concealed networks, thus making RDS an appropriate sampling method. Risk of ineligible persons attempting to participate due to the associated double incentive was noted, and using student identification documents as part of eligibility screening recommended. TSMSM also perceived RDS to be an acceptable strategy based on their large social network sizes (10-40), and the trust amongst themselves. TSMSM were concerned about participating due to the risk of being outed as MSM, seeing as same sex behavior is criminalized in Kenya, and hence emphasized that researchers needed to assure them of their confidentiality, and include MSM as part of the study team to encourage participation. TSMSM suggested coupons should indicate value of reimbursement, be pocket-sized and placed in an envelope to avoid loss, and provide directions to and contacts of the survey site for easy access.

**Conclusion:** RDS was perceived as both an appropriate and acceptable sampling method. Anticipated challenges of RDS implementation were highlighted, and possible solutions to these challenges suggested.

## Introduction

Globally, the HIV epidemic remains poorly defined among young MSM (YMSM) aged 15 – 24 years. There is minimal data on population size estimates of YMSM, their HIV rates as well as risk and protective factors. This is partially attributed to lack of research and surveillance, and the difficulty of reaching YMSM with services(1), especially in countries, like Kenya, where same-sex behavior continues to be criminalized(2). Additionally, YMSM are vulnerable to HIV due to widespread discrimination and stigmatization, homophobia and violence, power imbalances in relationships and alienation from family and friends(3). Largely due to these factors, YMSM are invariably at disproportionate risk of acquiring HIV/STI than young heterosexual males or older MSM(4).

During the transition period from adolescence to adulthood and a parallel shift from secondary to tertiary educational institutions, a majority of the students find themselves in an environment with limited direct supervision of their behavior and increased freedom and opportunities to engage in behavioral risks such as alcohol use and casual sex(5). For tertiary student MSM (TSMSM), this involves a sudden exposure to peers with similar sexual orientation, increased freedom and autonomy in young adulthood, and more socializing opportunities(6). Subsequently, they may engage in sex more frequently(7), and some might have unprotected anal intercourse more regularly(8), hence increasing their risk for HIV/STI infection. With most tertiary students in Kenya owning smart phones(9), TSMSM like other MSM may also use geosocial applications such as Grindr® to seek more sex partners for immediate sexual gratification, without paying attention to the accompanying risk of HIV/STI infection (10).

Even in countries where same-sex behavior is legal and data is available, TSMSM have been found to engage in high risk behavior that increases their susceptibility to HIV/STI. A systematic review and meta-analysis of studies published between 2003 and 2016 in China found out that TSMSM engaged in sex work, group sex, sex with female partners, drug use and seeking casual sex partners online(11). In the USA, TSMSM have been shown to have complex and expansive sexual networks within which HIV/STI may circulate widely and magnify epidemics(12). In South Africa, a behavioral survey among TSMSM showed a high prevalence of various risk factors for HIV transmission among TSMSM, including: high partner turnover, concurrent sexual partners, presence of STI, early sexual debut, having female sex partners, forced sex experiences, inconsistent condom usage, and alcohol and drug use(13).

In Kenya, same-sex behavior is criminalized(2). This may further amplify the impact of behavioral risk taking on HIV/STI transmission, over and above what has been observed among TSMSM in countries where same-sex behavior is legal(11–13). A literature review of TSMSM in Kenya yielded no information on the prevalence of HIV/STI nor risk behaviors in this potentially at-risk sub-group of MSM. There is a need to conduct research to better understand the epidemic in this neglected sub-population and hence inform the form and design of interventions to combat the HIV/STI syndemic in this population.

Respondent-driven sampling (RDS) has become a popular method to assess HIV/STI and risk behaviors in hidden or hard-to-reach populations such as MSM. RDS is both a recruitment and statistical method that utilizes “snowball sampling”, where individuals recruit those they know, the recruited individuals in turn recruit those they know and so on until an adequate sample is reached. This recruitment is combined with a mathematical model that weights the sample to compensate for the non-random sampling(14). Using RDS makes it possible to draw statistically valid samples from populations for whom there is no sampling frame, and provide unbiased population estimates and confidence intervals of disease prevalence and risk behaviors such as unprotected anal intercourse among MSM(15). Recruitment begins with ‘seeds’ purposely selected from the target population and given at most three ‘coupons’ to recruit their peers into the study. Participants who are recruited by the ‘seeds’ form the first ‘wave’ of recruitment. Participants in the first ‘wave’ are then issued with ‘coupons’ to recruit the second ‘wave’ of participants. This process goes on until the calculated sample size is reached. To encourage recruitment, participants are given an incentive to complete the survey (primary incentive) and to recruit their peers (secondary incentive)(16).

Previous studies that investigated HIV/STI in TSMSM recruited participants through either convenience sampling(17–19), or conducted studies among the general MSM population then drew a subset of TSMSM from this sample(20), or conducted studies among the general tertiary student population then drew a subset of TSMSM from this sample(7,13,21).

Formative research helps researchers and public health practitioners reach their target population and define that population’s characteristics that are pertinent to a particular public health issue of interest(22). Assessing appropriateness and acceptability is important so as to gauge the suitability of a particular method for a given population, and the agreeability of that method to a range of stakeholders, respectively(23). We conducted formative research to investigate these two outcomes for RDS as a recruitment strategy for TSMSM in a proposed HIV/STI bio-behavioral survey(24); and to identify potential challenges that would be faced with the use of this method, as well as possible solutions to these pitfalls. The study was done in Nairobi, the capital of Kenya.

## Materials and methods

### Theoretical framework and reporting

The study was underpinned by the content analysis theoretical framework, where data is systematically organized into a structured format(25). Reporting of the study was done following the consolidated criteria for reporting qualitative studies (COREQ)(26). [See S1 file].

### Site

The study was conducted in Nairobi, the capital city of Kenya and home to more than 50 campuses of various universities and, technical and vocational education training (TVET) institutions.

### Participants

Service providers were purposely selected from community-based organizations (CBOs that provide MSM-exclusive health services (n=2) and from a non-governmental organization (NGO) that provides MSM-friendly health services (n=1). The three organizations were selected purposely, first, because they are the main providers that offer MSM-exclusive/friendly health services in Nairobi, and are popular with both YMSM and TSMSM. Secondly, we sought to obtain information on how to best reach different typologies of TSMSM, with the ‘more out’ TSMSM assumed to be more likely to use the MSM-exclusive services and those ‘less out’ the non-exclusive but MSM-friendly services. Thirdly, the three organizations are conveniently located, with the NGOs being within and the CBOs located about 5 km from the central business district, hence easily accessible by TSMSM.

TSMSM were purposively recruited with the help of the service providers at the three facilities. After completing the interviews, the service providers were given the telephone contact of one member of the study team, and asked to request TSMSM known to them to contact the member of the study team regarding possible participation in the formative research. The service providers were given the ideal qualities of the TSMSM the study team was interested in speaking to, including: peer leaders with a social network size of at least nine TSMSM, interested in research and working with the TSMSM community. Consequently, contact was established with 16 TSMSM, out of which 13 were interviewed. Of the three who were not interviewed, one had already completed university education, one was not studying in Nairobi and the third one was unable to create time for the interview.

### Data collection

Semi-structured interview guides were obtained from the Integrated HIV Bio-behavioural Surveillance Toolbox of the University of California, San Francisco(27), and modified to fit the purpose of the current study [see S2 file and S3 file]. All the interviews were conducted by the first author, who introduced himself to the participants and provided them with detailed information about the study, including the key components of the RDS method. Interviews with service providers consisted of five major domains, namely: service providers’ roles in their respective organization, type of services offered by the organizations, description of the TSMSM community and their networks, appropriateness and acceptability of using RDS to recruit participants for the HIV/STI bio-behavioral survey, and challenges expected with the use of RDS and proposed solutions to these challenges. Interviews with TSMSM consisted of three major domains, namely: their social networks, appropriateness and acceptability of using RDS to recruit participants for the planned HIV/STI bio-behavioral survey, and challenges expected with the use of RDS and proposed solutions to these challenges. The network size was estimated by asking the question: *‘How many TSMSM do you know by name, and they know you by name, and they live in and around the city of Nairobi?’*. Demographic information was also collected using a brief self-administered questionnaire on REDCap (Vanderbilt University).

Interviews with the service providers were held at their respective work places. Interviews with the TSMSM were held in a private room at the integrated counselling and education center of University of Nairobi health services department. Only the interviewer and one participant were present at the time of every interview. Interviews with the service providers lasted an average of 40 minutes, whereas those with TSMSM took an average of 35 minutes. For the interviews with TSMSM, saturation was reached by the 10^th^ participant. All interviews were held between September and October 2020.

The interviewer took notes during all the interviews. All interviews were audio-recorded, transcribed and translated to English in the few instances where Swahili was used. The interviewer then read the transcripts as he listened to the audio-recordings to ensure that the transcripts and translations if any, were accurate.

### Data analysis

Data in form of the transcripts was managed using NVivo software version 11 (QSR International). A thematic framework approach was adopted. Analysis was done both deductively and inductively, using themes set a priori and coding emerging ones, respectively. Themes were supported with verbatim excerpts from the transcripts. Coding was done independently by two members of the study team, who then compared codes for agreement and decided whether to merge some codes, get rid of others or come up with new ones.

### Trustworthiness of the data

The trustworthiness of the data was premised on the following criteria: credibility, transferability, dependability and confirmability(28,29). To ensure credibility, the study used well-established methods for qualitative investigation. Prior to data collection, the first author familiarized himself with the setup and roles of the participating CBOs/NGO by reading content posted on the organizations’ respective websites. During data collection, iterative questioning was deployed by rephrasing previously asked questions to gain deeper understanding of and confirm the information provided by participants at the first instance of questioning. Member checking was done “on the spot” by paraphrasing and summarizing what the participants said at intervals and at the end of the interviews, respectively. For transferability, details about the study participants, study design, data collection procedures and analysis have been made available to help the comprehension of the findings, and comparison/contrast of these findings with those of similar studies. To address dependability, the study processes have been reported in sufficient detail to enable future researchers reproduce the study, even if not necessarily to obtain similar results. For confirmability, individual interviews were used due to the sensitivity of the study topic occasioned by criminalization and marginalization of same-sex behavior in Kenya. Confirmability was also established by supporting the findings with verbatim quotes from the participants.

### Ethics statement

The study protocol was reviewed and approved by University of the Witwatersrand Human Research Ethics Committee-Medical (Reference number: M200215) and University of Nairobi-Kenyatta National Hospital Ethics and Research Committee (Reference number: P990/12/2019). The study also received a letter of support from the Key Populations Program of the National AIDS and STI Control Program (NASCOP), Ministry of Health, Government of Kenya. Participants provided written informed consent before taking part in the study.

## Results

### Participants’ characteristics

The service providers who participated from the CBOs were both male, in their 30’s and identified as gay. The service provider from the NGO was female, in her 40’s and identified as heterosexual. All the providers had college degrees. All the service providers had dual roles at their respective organizations. The service provider at the first CBO worked as a pre-exposure prophylaxis (PrEP) program officer/research coordinator, the one at the second CBO as a community mapping officer/peer educators’ supervisor and the one at the NGO as a clinician/head of the clinic. These service providers were selected on the basis of their day-today interaction with members of the MSM population, and the assumption that they would have rich information on the TSMSM sub-population who were part of their clientele. All the three organizations offered HIV/STI prevention, treatment and care services. Additionally, the two CBOs offered other services including: community outreaches, economic empowerment of, and provision of legal aid to the MSM communities they served. On the other hand, the NGO offered care to victims of gender-based violence, and sexual and reproductive health services to other key populations such as female sex workers. The service providers referred 16 TSMSM to the research team, out of which 13 participated in the formative research (n=13).

Demographic characteristics of the TSMSM participants are presented in **Table 1**. More than three-quarters (76.9%) were 21-24 years of age. All participants were undergraduate students in their first (23.1%), second (38.5%) or third (38.5%) years of study. A majority (69.2 %) identified as homosexual or gay and the rest as bisexual.

**Table 1:** Demographic characteristics of TSMSM participants (n=13)

| **Variable** | **n (%)** |
| --- | --- |
| **Age** |  |
| 18-20 | 3 (23.1) |
| 21-24 | 10 (76.9) |
| **Level of education** |  |
| Undergraduate | 13 (100) |
| Year 1 | 3 (23.1) |
| Year 2 | 5 (38.5) |
| Year 3 | 5 (38.5) |
| **Type of institution** |  |
| University | 11 (84.6) |
| *TVET/College | 2 (15.4) |
| **Ownership of institution** |  |
| Public | 9 (69.2) |
| Private | 4 (30.8) |
| **Field of study** |  |
| Arts and humanities | 3 (23.1) |
| Business and management | 3 (23.1) |
| Engineering and technology | 3 (23.1) |
| Health sciences | 2 (15.4) |
| Social sciences | 2 (15.4) |
| **Residence** |  |
| Rented (apartment/flat/hostel outside college/university) | 8 (61.5) |
| At home with family | 5 (38.5) |
| **Main source of income** |  |
| Part time employment | 8 (61.5) |
| Parents/guardians | 5 (38.5) |
| **Network size** |  |
| 10 – 24 | 6 (46.2) |
| 25 – 40 | 7 (53.8) |
| **Sexual orientation** |  |
| Homosexual or gay | 9 (69.2) |
| Bisexual | 4 (30.8) |

\*TVET: Technical and vocational education training institution

The following explores main themes from the analysis of the interviews with both the service providers and TSMSM. Themes are described and supported with illustrative quotes from the interviews. Quotes are attributed to participants using pseudonyms and role (service provider or student).

### Social networks of TSMSM

A key theme that emerged from the interviews is that TSMSM constituted a large and well networked sub-group of the MSM community. TSMSM formed networks among themselves, as well as with other sub-groups of MSM. TSMSM had regular face-to-face interactions with their peers from the same institution and those from different institutions. However, since the outbreak of COVID-19, these interactions had been hampered by government-imposed restrictions to reduce risk of disease transmission. Consequently, TSMSM mainly interacted through social media applications such as Facebook, WhatsApp, Twitter and Instagram.

> *“In the student MSM community, what happens is that there is some sort of invisible network that is hidden but it is very large and well connected*.*”* – Justin, service provider
>
> *“I know about 30 to 40 people (other TSMSM in Nairobi). I have seen about 10 (in the last 30 days) … due to the COVID situation, so most people travelled (out of Nairobi) … most of them are the people we study with, like in the same institution… but I also know others from other universities*.*” –* Kyle, student
>
> *“Twenty-five to thirty (number of TSMSM known). Due to the corona (COVID-19), I have seen about 12… we are used to each other… we always chat through WhatsApp, Facebook and Twitter sometimes… even through Instagram*.*” –* Caleb, student

TSMSM exhibited strong ties with their peers (intra-group ties), through which they found safe spaces to socialize such as night clubs and house parties. The house parties offered opportunities to engage in risky behaviors such as use of alcohol and casual sex. In spaces where there were other MSM such as night clubs, TSMSM tended to interact and socialize with each other, staying away from other MSM perceived to have lower educational attainments.

> *“They (TSMSM) love parties… like on Fridays, they have house parties among themselves… so they really interact among themselves so much, then they get drunk and have sex*.*”* – Nicole, service provider
>
> *“Most of them (MSM) group themselves into clusters, whereby even if you get into a club, you will find them like the tertiary ones (students) are seated at one corner, and the lower end ones (with lower education levels) at the other corner*.*”* – Mike, service provider

TSMSM also interacted with other sub-groups of MSM. TSMSM were a bridging sub-population between well-off, older MSM and less well-off MSM within their age-group, and engaged in transactional sex, receiving money from the former and in turn giving money to the later.

> *“MSM students have sponsors (older and well-off MSM) … because they want to have a flashy lifestyle, they want to have money, so they will have sponsors to sponsor their lifestyles*.*”* – Nicole, service provider
>
> *“Tertiary MSMs, they have sex with those guys with money so that they can cater for their needs. Once (after) they are from that upper end, they will come into the clubs*… *they try to mingle and buy sex from the lower end partners*.*” –* Mike, service provider

### Appropriateness and acceptability of using RDS to recruit TSMSM in a prospective HIV/STI survey

Participants felt that RDS would be an appropriate and acceptable method of recruiting TSMSM to the prospective HIV/STI survey. TSMSM regarded RDS as an appropriate recruitment strategy due to the hidden but networked nature, and the trust that existed among members of the TSMSM community. The perceived trust among peers would enable them reach each other better, as compared to the research team trying to reach the TSMSM directly.

> *“It (RDS) would work best and I think it is one of the recommended methods especially MSM being hidden… of course they will tend to trust information coming from their peers and this group of MSM is actually very highly networked”* – Jay, student
>
> *“I think it (RDS) would work very well because it’s easier for me to refer someone than you going out there to look for a guy to participate… they (my friends) would believe my word of mouth so it’s convincing enough*.*”* – Trevor, student

Service providers noted that they were already known and trusted by TSMSM, and this existing relationship would make TSMSM referred by the service providers more likely to agree to participate in the prospective survey.

> *“If you are given something by a friend of yours, it is easy to accept it… but if you are given by a stranger, that is when challenges come in… because you (researchers) are already embedding yourselves in the existing networks, then it is okay because of the already existing trust between us (service providers) and the students*.*”* – Justin, service provider

The proposed double reimbursement for first participating and then getting a friend to participate in the prospective survey, also made RDS popular. The first reimbursement would be used to cater for expenses such as fare to get to the survey site, and the second one would motivate peers to bring in their friends to take part in the prospective survey.

> *“It (RDS) will be good because of the reimbursement… you are using your transport from where you live to the place where you are going to get the service or the research so it will be a good thing. Once you’ve told someone that the moment you bring someone there is something for you it’s not that hard (for a friend to agree to participate)*.*”* – Danny, student
>
> *“I think since there is a small reimbursement, I think that would motivate people to really recruit their friends to come for the study*.*”* – Kyle, student

### RDS anticipated challenges and proposed solutions

Participants anticipated that RDS implementation would face some challenges, and offered probable ways of mitigating these challenges. Key among the anticipated challenges was that if the survey was associated with CBOs that serve sex workers, then TSMSM would not be willing to take part in the survey, since they would not want to be associated with sex work. It was proposed that the researchers present themselves to possible participants as independent from these CBOs so as to encourage participation.

> *“The only challenge will be if this (the survey) is associated with some organizations it might discourage some people from participating. You find this group (TSMSM) sometimes doesn’t want to be associated with some community clinics because they have a reputation of serving sex workers… I guess I like that you presented yourself as an independent researcher from an academic institution, that is straightforward… people will feel free to participate*.*”* – Jay, student

Another foreseen challenge was the possible loss of recruitment coupons. Participants suggested various ways of handling this, including: indicating the incentives on the coupons so that TSMSM would be motivated to keep the coupons safely, having coupons in a small size that would easily fit in the pocket, and placing coupons in an envelope.

> *“They (coupons) get lost a lot, someone just presents himself without it. If they are communicating the incentive directly, then it works… someone will be able to store it properly*.*”* – Jay, student
>
> *“The challenge I was thinking of is when it (the coupon) gets lost… but if it will be in a smaller size, a pocketable size it is not easy to lose it*.*”* – Frank, student
>
> *“The person can lose the coupon. If it can be sealed in an envelope well, then it will be easy for them to handle*.*”* – Caleb, student

It was also noted that some TSMSM would be unwilling to participate if they were not assured about confidentiality with regard to their same-sex behavior. To encourage TSMSM recruitment, it was suggested that during consenting, TSMSM be provided with adequate information on how their privacy and confidentiality would be guaranteed, and assurance that it was safe to participate in the survey.

> *“I believe with detailed information, if one got the information correctly, maybe the willingness (to participate) would be much better*.*”* – Kyle, student
>
> *“Most of them (MSM), I think you need to assure them of the confidentiality in the interview. That is the thing that most MSM fear. Maybe they will be mentioned out there, or if they are students like at the university, they will be discriminated*.*” –* Pitts, student

To further encourage recruitment, it was suggested that part of the survey team be MSM since TSMSM would be more comfortable opening up to other MSM.

> *“Due to the sensitivity of what you’re dealing with, only some of my friends would agree to come for the study… most of them are ok opening up to fellow members (MSM) but when it comes to somebody else they don’t open up. So it is good to have other MSM working with you*.*”* – Perry, student

Participants also predicted that TSMSM would encounter a twofold challenge in getting to the survey site – knowing the location of the survey site and having the means to get to the survey site. To allay this challenge, participants proposed that the survey team should consider: giving directions to the survey site by sharing a location pin on smart phones, providing a telephone number TSMSM could call if they needed clarifications about the location of the survey site, and reimbursing participants with money to cover transport costs.

> *“In terms of direction, if someone has a smartphone you can send them the location pin [GPS coordinates] of the study office. It could work… instead of walking around asking from people… and include phone details just for inquiries*.*” –* Andy, student
>
> *“You might find someone does not have the transport to get to town. If there could be a way they could be sorted out when they come, they get reimbursed the amount… I think it would work. It would give someone motivation to come and participate*.*”* – Andy, student

From the service providers’ point of view, it was noted that due to the proposed incentives in the prospective survey, there was a likelihood of recruiting MSM who were not TSMSM and/or men who were not MSM. To mitigate this to some extent, it was recommended that the research team could ask to see student identification cards and compare the student’s facial appearance with the image on the identification cards.

> *“When someone wants to get the incentive and does not have friends who are students, you are likely to be brought people who are coached… you are also likely to be brought people who are not MSM who are also coached*.*”* – Nicole, service provider
>
> *“To participate, one comes with his school ID… but now, one can borrow an ID… it is still not 100% viable (foolproof) but it tries to limit number of non-students participating*.*”* – Nicole, service provider

Finally, carrying out the prospective survey when tertiary learning institutions were closed due to COVID-19 restrictions, or when students were having examinations, would interfere with recruitment of participants. It was proposed that the prospective survey be conducted at a time when COVID-19 restrictions would have eased, and students were not sitting examinations.

> *“For a student it depends on whether he is currently in college or at home because of corona (COVID-19) … and also, are you giving these coupons during exams time? Of course no one would come if they have exams. And you know students at university, they only read during exams time and therefore they would not bring back the coupons during that time*.*”* – Justin, service provider
>
> *“It is better if you do the study when universities re-open (after COVID-19 restrictions have eased) and also when the students are not having exams*.*”* – Mike, service provider

## Discussion

We assessed whether RDS was an appropriate and acceptable strategy for recruiting young TSMSM in a HIV/STI bio-behavioral survey. We also assessed barriers and facilitators as well as challenges anticipated with the use of this method, and possible solutions to these challenges. RDS relies on certain assumptions that need to be met for RDS to be considered an appropriate sampling method for a particular population. These assumptions are: (1) the target population is sufficiently networked to sustain recruitment through the waves and eventually reach the desired sample size, (2) peer relationships are reciprocal (*a* knows and can recruit *b, b* knows and can recruit *a*), (3) recruitment within an individual’s network is random and (4) sampling occurs with replacement so that the set of available recruits is not depleted(30). Our findings demonstrate that the first two assumptions were met for the TSMSM population. To begin with, the TSMSM population was well networked with members exhibiting close social ties and having large network sizes. Secondly, members of the TSMSM population formed reciprocal relationships characterized by trust between each other. Based on the fulfillment of these two assumptions, RDS would thus be an appropriate recruitment strategy for TSMSM. Besides the trust that existed between peers, RDS was also acceptable to TSMSM due to the proposed double incentives, and the discrete approach the method uses.

Our target population of TSMSM was young and socially marginalized. Other studies have shown that, to varying degrees, RDS is an appropriate and acceptable strategy for recruiting university students(31), sexual and gender minority youth, including young MSM(32), and young adult non-medical users of pharmaceutical opioids(33) into research. However, other studies have found that RDS may be an inappropriate strategy for recruiting socially marginalized populations such as urban young MSM(34) and young illicit drug users(35), mainly due to members of the target populations having small network sizes and insufficient ties within their networks, thus the inability to sustain recruitment. Notably, the studies highlighted here used data that was collected after the RDS surveys had been completed. To the best of our knowledge, this is the first study to use formative research to assess the appropriateness and acceptability of using RDS to recruit young TSMSM for HIV/STI research in sub-Saharan Africa. As in other studies(36–38), this formative research provided us with useful information on the challenges that would be expected with the use of RDS, and probable solutions to these challenges. Being aware of these challenges is important to research teams, since the teams are then able to put in place measures to address these challenges prior to implementing RDS surveys.

Our study had some limitations. By nature, formative research is not definitive, neither is it designed to test hypotheses. Rather, formative research can help inform whether RDS is an appropriate and acceptable strategy for the target population, as well as contribute to the RDS survey planning process. We cannot therefore say for sure that based on these findings that an RDS survey among TSMSM would be successful. We would have to conduct the RDS survey itself, then evaluate its outcomes and consider the value of the formative research to the RDS survey. Another limitation of this study is that we interviewed a small convenient sample of service providers. However, we felt that with this sample, given the dual roles each of the providers had at their respective organizations, and the day-to-day interactions the providers had with MSM, we would be able to obtain sufficient information to meet the objectives of our study. Additionally, the three organizations from which the participants were sampled from are the primary providers of MSM exclusive/friendly services and are highly rated by MSM. As well, the study was conducted at a time when there were government-imposed COVID-19 restrictions, with most staff in these organizations working from home thus limiting their availability and accessibility to participate in the study. Finally, the study was conducted in the capital city which is expected to be more liberal than other smaller cities or rural settings in the country, thus limiting the generalizability of our findings. Nonetheless, these findings demonstrate the appropriateness and acceptability of RDS among TSMSM, in addition to bringing to light the anticipated challenges of using RDS and offering recommendations on how these challenges could be addressed prior to implementing the RDS survey.

## Conclusions

TSMSM had large network sizes and strong ties within the networks, properties considered to be appropriate to sustain recruitment through the RDS waves. The RDS method was acceptable to TSMSM due to the trust that existed amongst them, hence making it easier for them to reach each other as compared to the researchers themselves attempting to reach TSMSM directly. The study also offered valuable lessons on the challenges that would be expected with the use of RDS in the TSMSM population, and proposed possible solutions to these challenges. Pre-assessing the suitability and agreeability of methods to reach young marginalized populations appears to be a practical way to begin to gain entry into these populations, ensuring that the views and voices of possible future participants are incorporated into the design of prospective studies.

## Supporting information

COREQ checklist

Interview guide for service providers

Interview guide for TSMSM

## Data Availability

All data produced in the present study are available upon reasonable request to the authors.

## Acknowledgements

The authors would like to thank the participants for agreeing to take part in the study. We also thank Joyce Kafu and Jacqueline Akinyi for transcribing the audio-recordings of the interviews. We are also grateful to Catherine Ngugi – Head NASCOP, and Helgar Musyoki – Key Populations Program Manager at NASCOP for supporting this study.

## Participants’ consent for publication

Obtained

