## Supplementary material for "“My friends would believe my word”: appropriateness and acceptability of respondent-driven sampling in recruiting young tertiary student men who have sex with men for HIV/STI research in Nairobi, Kenya": COREQ checklist

| **Number** | **Where reported in the manuscript** |
| --- | --- |
| **Domain 1: Research team and reflexivity** | |
| ***Personal characteristics*** | |
| 1. Interviewer | Data collection |
| 2. Credentials | Title page (link to ORCiD) |
| 3. Occupation | Title page (link to ORCiD) |
| 4. Gender | Title page (link to ORCiD) |
| 5. Experience and training | Title page (link to ORCiD) |
| ***Relationship with participants*** | |
| 6. Relationship established | Data collection |
| 7. Participant knowledge of the interviewer | Data collection |
| 8. Interviewer characteristics | Data collection |
| **Domain 2: Study design** | |
| ***Theoretical framework*** | |
| 9. Methodological orientation and theory | Theoretical framework |
| ***Participant selection*** | |
| 10. Sampling | Participants |
| 11. Method of approach | Participants |
| 12. Sample size | Participants |
| 13. Non-participation | Participants |
| ***Setting*** | |
| 14. Setting of data collection | Data collection |
| 15. Presence of non-participants | Data collection |
| 16. Description of sample | Participants and results |
| ***Data collection*** | |
| 17. Interview guide | Data collection |
| 18. Repeat interviews | N/A |
| 19. Audio/visual recording | Data collection |
| 20. Field notes | Data collection |
| 21. Duration | Data collection |
| 22. Data saturation | Data collection |
| 23. Transcripts returned | N/A |
| **Domain 3: Analysis and findings** | |
| ***Data analysis*** | |
| 24. Number of data coders | Data analysis |
| 25. Description of the coding tree | Results |
| 26. Derivation of themes | Data analysis |
| 27. Software | Data analysis |
| 28. Participant checking | Trustworthiness of the data |
| ***Reporting*** | |
| 29. Quotations presented | Results |
| 30. Data and findings consistent | Results |
| 31. Clarity of major themes | Results |
| 32. Clarity of minor themes | N/A |
