## Supplementary material for "“My friends would believe my word”: appropriateness and acceptability of respondent-driven sampling in recruiting young tertiary student men who have sex with men for HIV/STI research in Nairobi, Kenya": Interview guide for service providers

**Guide for interviews with service providers:**

**Exploring the appropriateness, acceptability and challenges of RDS in recruiting TSMSM into a prospective HIV/STI bio-behavioral survey**

**Introduction:** I would like to thank you for agreeing to participate in this interview. We are going to talk about a study we are planning to carry out on HIV and STI among TSMSM. We would like to hear your views about the methods we are thinking of using in this study so that you can let us know if the methods are suitable or not, and what we can do to improve them. Please feel free to engage in this conversation.

| **Themes** | **Questions** | **Probes/prompts** | **Notes** |
| --- | --- | --- | --- |
| I would like to start by asking you about what you do in this organization: | | | |
| Role in the organization | Tell me about your role in this organization: | What do you do on a day-to-day basis?  What else do you do apart from what you have told me? |  |
|  | What type of contact do you have with MSM clients? | How often do you work with MSM? Every day? Occasionally? Rarely?  Are some of the MSM you see tertiary/college/university students?  If so, how often do you work with these TSMSM? |  |
| Now, I would like to ask you about the type of services offered by your organization: | | | |
| Services offered by the organization | What type of services does your organization provide? | HIV/STI awareness? Prevention? Treatment?  Psychosocial support? |  |
|  | Apart from what your organization offers, what other services exist for MSM in and around Nairobi? | HIV/STI awareness? Prevention? Treatment?  Psychosocial support?  Which organizations offer these services? |  |
|  | Who are the key population for your services? | MSM? Sex workers?  Injecting drug users?  Adolescents and young women? |  |
|  | Approximately how many MSM did your organization serve this past year (last 12 months)? | Of these, approximately how many/ what proportion were TSMSM?  In your day-to-day record keeping, do you have a way of documenting an MSM client’s level of education? |  |
| Now I would like you to help me understand the MSM community/networks in and around Nairobi: | | | |
| MSM community and networks | Think about MSM in and around Nairobi. How can you describe them? | Could you describe for me the different subgroups of MSM?  Do they differ by: Age; Education; Income; Ethnicity; Gender identity; Sexual preferences; Other?  How much interaction or contact is there between the sub-groups?  How do TSMSM interact with the different sub-groups that you just described? |  |
| Now, I would like to describe to you how we plan to recruit TSMSM into the HIV/STI study. I will then ask you some questions that will help us plan the study better.  We are thinking of choosing a few TSMSM who are well connected and regarded in the TSMSM community. We will give them some invitation coupons that they could give to their peers to invite them to participate in the study. For every friend/peer they recruit into the survey, they will receive a cash incentive on top of the cash incentive they receive for participating themselves. The participants that come in will then be given coupons that they can use to recruit their friends/peers. The process will continue until we reach our sample size of 250 TSMSM. *[SHOW participant a sample of the invitation coupon and explain how it is used*]. | | | |
| Appropriateness and acceptability of RDS | How well do you think this method would work in recruiting TSMSM into the study? | Is it a good method? Why?  Is it a bad method? Why? |  |
|  | Do you think TSMSM would be willing to invite other TSMSM using the invitation coupons? | What would make them to be willing?  What would make them to be unwilling? |  |
|  | After receiving an invitation coupon, how long do you think TSMSM would take to come in and participate in the study? | 1,2,3 days? A week? 2 weeks? 1 month?  What factors would influence how fast TSMSM come to participate after receiving the coupon? COVID-19 restrictions? Schedule of classes? Examinations? Transport to the study site? |  |
|  | Approximately how long do you think it would take us to recruit 250 TSMSM into the study? | 1,2 or 3 months? 6 months? 1 year?  What factors would influence how long it would take us to achieve our sample size?  COVID-19? Incentives? Schedule of classes? Examinations? |  |
| RDS challenges and proposed solutions | What challenges are we likely to encounter when using this method of recruitment? | Failure to get participants?  Participants who are not MSM?  MSM who are not tertiary students? |  |
|  | How do you suggest we could address these challenges? | Extra incentives?  Screening to ensure participants are MSM?  Use of student IDS? |  |

**Conclusion:** Do you have any questions about the study before we end? Is there anything you would like to add to our discussion? [*TAKE time to address all questions and concerns*].

Thank you very much for taking time to talk to us. The information you have provided us with is important and will help us in planning for the study.
