## Supplementary material for "“My friends would believe my word”: appropriateness and acceptability of respondent-driven sampling in recruiting young tertiary student men who have sex with men for HIV/STI research in Nairobi, Kenya": Interview guide for TSMSM

**Guide for interviews with TSMSM:**

**Exploring the appropriateness, acceptability and challenges of RDS in recruiting TSMSM into a prospective HIV/STI bio-behavioral survey**

**Introduction:** I would like to thank you for agreeing to participate in this interview. We are going to talk about a study we are planning to carry out on HIV and STI among TSMSM. We would like to hear your views about the methods we are thinking of using in this study so that you can let us know if the methods are suitable or not, and what we can do to improve them. Please feel free to engage in this conversation.

| **Themes** | **Questions** | **Probes/prompts** | **Notes** |
| --- | --- | --- | --- |
| I would like to start by asking you about your friends. | | | |
| Social networks | Tell me about your closest friends: | Are your friends also TSMSM?  Where do they study? Same institution as you or different one(s)?  Do you have friends who are not TSMSM? How would you describe them? |  |
|  | How many TSMSM do you know by name, and they know you by name, and they live and study in and around the city of Nairobi? | How many of these have you seen in the past one month?  How about in the past one week?  Have the COVID-19 restrictions affected how often you see your friends who are TSMSM? Do you see them more or less often? |  |
|  | How easy is it for you to contact your TSMSM friends? | Easy? Difficult? Not easy not difficult? |  |
|  | How do you contact your TSMSM friends? | Phone calls? Text messages? Social media (WhatsApp, Telegram, Instagram, Twitter)? |  |
| Now, I would like to describe to you how we plan to recruit TSMSM into the HIV/STI study. I will then ask you some questions that will help us plan the study better.  We are thinking of choosing a few TSMSM who are well connected and regarded in the TSMSM community. We will give them some invitation coupons that they could give to their peers to invite them to participate in the study. For every friend/peer they recruit into the survey, they will receive a cash incentive on top of the cash incentive they receive for participating themselves. The participants that come in will then be given coupons that they can use to recruit their friends/peers. The process will continue until we reach our sample size of 250 TSMSM. *[SHOW participant a sample of the invitation coupon and explain how it is used*]. | | | |
| Appropriateness and acceptability of RDS | How well do you think this method would work in recruiting TSMSM into the study? | Is it a good method? Why?  Is it a bad method? Why? |  |
|  | If you are selected and given say 3 coupons, how long would it take you to get 3 of your friends/peers to give them the coupons? | 1,2,3 days? A week? 2 weeks? 1 month? |  |
|  | After giving a friend an invitation coupon, how long do you think they would take to come in and participate in the study? | 1,2,3 days? A week? 2 weeks? 1 month?  What factors would influence how fast a friend comes to participate after receiving the coupon? COVID-19 restrictions? Schedule of classes? Examinations? Transport to the study site? |  |
|  | Do you think your friends would be willing to recruit their friends using such coupons? | What would make them to be willing?  What would make them to be unwilling? |  |
|  | Approximately how long do you think it would take us to recruit 250 TSMSM into the study? | 1,2 or 3 months? 6 months? 1 year?  What factors would influence how long it would take us to achieve our sample size?  COVID-19? Incentives? Schedule of classes? Examinations? |  |
| RDS challenges and proposed solutions | What challenges are we likely to encounter when using this method of recruitment? | Failure to get participants?  Participants who are not MSM?  MSM who are not tertiary students? |  |
|  | How do you suggest we could address these challenges? | Extra incentives?  Screening to ensure participants are MSM?  Use of student IDS? |  |

**Conclusion:** Do you have any questions about the study before we end? Is there anything you would like to add to our discussion? [*TAKE time to address all questions and concerns*].

Thank you very much for taking time to talk to us. The information you have provided us with is important and will help us in planning for the study.
